# Serum Thyroid-Stimulating Hormone and 25-Hydroxycholecalciferol Levels in Children with Autism Spectrum Disorder and Intellectual Disability in Northern India: A Case-Control Study

**DOI:** 10.1101/2022.12.23.22283891

**Authors:** Shivam Singh, Pragati Basera, Preeti Agarwal, Amit Arya, Wahid Ali, Kopal Rohatgi, Ahmad Ozair

**Affiliations:** Faculty of Medicine, King George’s Medical University, Lucknow, Uttar Pradesh, India; Department of Pathology, King George’s Medical University, Lucknow, Uttar Pradesh, India; Department of Psychiatry, King George’s Medical University, Lucknow, Uttar Pradesh, India; Miami Cancer Institute, Baptist Health South Florida, Miami, FL

**Keywords:** autism, vitamin D, disability, subclinical hypothyroidism, resource-limited setting

## Abstract

**Background:** Autism spectrum disorder (ASD) and intellectual disability (ID) affect 2-3% of the global population with both conditions having unclear pathophysiology. Little data exists from South Asia examining the association between these conditions and the disturbances in thyroid profile and vitamin-D status.

**Objective:** This study sought to characterize the association between these conditions and serum thyroid-stimulating hormone (TSH) and 25-hydroxycholecalciferol (25(OH)D3) levels, from a resource-limited setting in India.

**Methods:** The present work was a prospective, multi-arm, case-control study conducted and reported in accordance with Strengthening the Reporting of Observational Studies in Epidemiology (STROBE) guidelines. Children with ASD, children with ID, and age-and-sex matched healthy controls, attending the outpatient clinics of pediatrics and psychiatry in Northern India were included. Primary outcomes were serum TSH and 25(OH)D3 levels, which were measured by chemiluminescent immunoassay and compared using ANOVA.

**Results:** A total of 45 children were included of which 15 had ASD, 30 with ID, and 30 were controls. There were 44 males and 31 females, with a mean age of 7.01±2.77 years. Mean±SD levels of 25(OH)D3 were significantly lower in ASD (9.53±4.93 IU/mL) and ID (14.39±5.99 IU/mL) compared to healthy controls (16.37±10.68 IU/mL) [p=0.032]. Mean±SD levels of TSH were similar in ASD (2.65±1.96), ID (2.47±2.03), and controls (2.19±1.42), with no significant difference [F-value=0.37; P=0.69]. 8% (N=6/75) of the participants had a raised TSH level.

**Conclusion:** In conclusion, children with ASD and ID have significantly lower vitamin D3 levels compared to healthy controls. High-quality randomized controlled trials are warranted to further investigate the therapeutic impact of early-life vitamin D supplementation in these indications.

**Lay Summary:** Autism spectrum disorder (ASD) and intellectual disability (ID) affect 2-3% of the global population with both conditions having multiple etiologies. Little data exists from South Asia examining the association between these conditions and the disturbances in thyroid profile and vitamin-D status. This study aimed to explore the association between these conditions and serum thyroid-stimulating hormone (TSH) and 25-hydroxycholecalciferol (25(OH)D3) levels, from a resource-limited setting in India. For this, 15 children with ASD, 30 with ID and 30 healthy controls were recruited. Their serum TSH and 25-hydroxycholecalciferol levels were measured and compared. It was found that children with ASD and ID have significantly lower vitamin D3 levels compared to healthy controls, while levels of serum TSH were similar. High-quality randomized controlled trials are needed to explore the therapeutic impact of early-life vitamin D supplementation in ASD and ID.

## 1. INTRODUCTION

Autism spectrum disorder (ASD) is a widely prevalent condition, with a WHO review estimating its global prevalence to be nearly 1% (Elsabbagh et al, 2012). More recent reviews indicate this prevalence to be closer to 1.5% in developed countries (Lord et al, 2018). The condition is estimated to afflict nearly 1 out of every 60 children, being more frequent in males. The reported prevalence of ASD has gradually increased over the past few decades given the steady improvements in public awareness, availability of management services, medical documentation, and changes in diagnostic criteria that favor the inclusion of milder cases without intellectual disability (ID) (Lord et al, 2018). ASD represents a significant economic burden, both due to the loss of productive individuals and the resources required to support their well-being. Genetic, environmental, neurologic, immunologic, and other factors have been found to be linked with the development of ASD, however, the precise etiology and pathogenesis yet remain unclear (Lord et al., 2018; Desoky et al., 2017).

Intellectual disability (ID), a condition with some overlap with ASD, is estimated to variably impact 2-3% of the population (Daily et al., 2000). ID is currently diagnosed when the intelligence quotient (IQ) of a subject, if obtained in appropriate settings, is below 70, according to the Diagnostic and Statistical Manual of Psychiatric Disorders, Fifth Edition (DSM-5) (American Psychiatric Association, 2013). Nearly half of the patients with ID have an unknown etiology apart from associated genetic factors, toxins, and psychosocial illness (Sadock et al., 2017).

Thyroid-related abnormalities have been found to be associated with neurodevelopmental illnesses, including ASD and ID (Segni, 2017, Prezioso et al., 2018, Hoshiko et al., 2011). Through their actions on regulatory genes involved in neurogenesis, thyroid hormones (TH) are essential for brain development (Rovet, 2014). It has been reported that nearly half of children with ASD suffer from some form of either clinical or subclinical thyroid dysfunction (Loyacono et al, 2020). Similarly, vitamin D-related abnormalities have been found to be associated with neurodevelopmental disorders, including ID and ASD. 1,25-dihydroxycholecalciferol (Vitamin D3) performs neuroprotective, immunomodulatory, and anti-inflammatory roles. Its neuroprotective role, in particular, is evident in its regulation of the seizure threshold, and maintenance of the T-regulatory cell population, amongst other effects. Reduction in vitamin D levels results in aberration of the global immune-modulatory role, for instance, the association between low Vitamin D3 levels and autoimmune thyroiditis (Mackawy et al., 2013). Patients with ID have been found to have nearly twice the rate of vitamin D deficiency compared to controls in the UK (Frighi et al, 2014).

However, most of the work regarding associations between (1) ASD and ID, and (2) aberrancies of thyroid hormones and vitamin D physiology, has come from high-income countries (HICs), with little contemporary data from low-and-middle-income countries (LMICs). Given that populations in LMICs have significant genetic and environmental differences, it, therefore, stands to reason that LMIC populations may or may not have these explicit associations reported from HIC populations, which we aimed to investigate. To the best of our knowledge, there do not exist any peer-reviewed publications exploring these associations in the Indian population.

Given that both thyroid hormones and vitamin D have been implicated in several neurodevelopmental illnesses with partially unclear mechanisms, this work, therefore, sought to look for the deficiencies of thyroid hormones and vitamin D in children with ASD and ID in a resource-limited setting in South Asia.

## 2. METHODS

### 2.1 Study Design

The present work was a prospective case-control study conducted and reported in accordance with the Strengthening the Reporting of Observational Studies in Epidemiology (STROBE) guidelines. The study was performed at the Departments of Psychiatry, Pediatrics, and Pathology in a tertiary-care, publicly funded institution in Northern India, with the study hypothesis, “hypothyroidism and vitamin D deficiency are more prevalent in children with ASD and ID”.

### 2.2 Study Participants

For this work, three groups were included from amongst children and adolescents, having clinically diagnosed ASD (Group A) and clinically diagnosed ID (Group B), and age-and sex-matched healthy controls (Group C). DSM-5 criteria were utilized for all diagnostic evaluations. The DSM-5 Text Revision (DSM-5-TR) given that they had not been published at the time of the data collection.

All participants were within the age group of 2-12 years. Assent from the participants and written informed consent from their guardians were obtained before enrolment. We excluded any individual from any of the case or control groups if there was 1) presence of medical/surgical/psychiatric illness that requires priority management, and 2) refusal of guardian”s informed consent.

### 2.3 Study Outcomes

The primary outcomes were levels of 25-hydroxycholecalciferol and thyroid-stimulating hormone (TSH), which were recorded on the ARCHITECT i1000SR immunoassay analyzer by chemiluminescent micro-particle immunoassay technique (Abbott Diagnostics, Abbott Park, Illinois, U.S.). Using serum 25-hydroxycholecalciferol, patients were classified as deficient and non-deficient using the updated cutoff of 12 ng/mL (30 nmol/mL), as recommended by the updated cutoff, recommended in the New England Journal of Medicine by the members of the Institute of Medicine (IOM) committee which had previously provided recommendations for daily intakes in 2010 (Manson et al, 2016, Shah et al, 2017, Rosen et al, 2012, Ross et al, 2011).

### 2.4 Data Collection

Eligible cases of ASD and ID attending child and adolescent psychiatry outpatient clinic (OPD) were prospectively screened and all consecutive cases fulfilling inclusion criteria were included. The control group was selected from Pediatrics OPD IQ assessment of all included cases with ASD and ID was performed. Similarly, a formal assessment of intellectual functioning for done for all included controls. Data on sociodemographic and clinical variables of cases and controls were recorded on semi-structured proforma. 5 ml of blood was collected by venipuncture and after serum separation, the sample was analyzed for levels of 25-hydroxycholecalciferol and TSH.

### 2.5 Statistical analysis

Statistical analysis was carried out in Statistical Package for the Social Sciences (SPSS) software version 16 (IBM SPSS Statistics for Windows, IBM Corp, Armonk, NY, US). Intergroup comparisons of categorical variables were performed using the Chi-square test, while serum levels of 25-hydroxycholecalciferol and TSH were compared through the Analysis of Variance (ANOVA) test. Statistical significance was set at a two-tailed p-value of 0.05.

### 2.6 Ethical Considerations

The work was approved by the institutional ethics committee (reference number 95^th^ ECM-IIB-IMR-S/P2) and conducted in accordance with the ethical guidelines of the Declaration of Helsinki. Informed consent had been collected from the parents of the children at the time of presentation for all further research activities. Confidentiality was strictly maintained through the use of an assigned identification number.

## 3. RESULTS

A total of 75 subjects were enrolled in the study, including 15 children with ASD (20%), 30 children with ID (40%), and 30 healthy controls [**Table 1**].

**Table 1:** Demographic and clinical variables of included individuals in the present study.

|  |  | N | Percentage |
| --- | --- | --- | --- |
| Participants | Cases with ASD | 15 | 20.0% |
|  | Cases with ID | 30 | 40.0% |
|  | Healthy Controls | 30 | 40.0% |
| Mean Age |  | 7.01±2.77 years |  |
| Sex | Male | 44 | 58.7% |
|  | Female | 31 | 41.3% |
| Serum 25(OH)-cholecalciferol | Non-Deficient | 52 | 69.33% |
|  | Deficient | 23 | 30.66% |
| Serum TSH | Normal | 69 | 92.0% |
|  | Raised | 6 | 8.0% |
| IQ Score Severity<br>In cases with ASD and ID | Normal | 6 | 13.3% |
|  | Mild | 22 | 48.9% |
|  | Moderate | 10 | 22.2% |
|  | Severe | 7 | 15.6% |
|  | Total | 45 | 100.0% |

Over 95% (N=72) of study participants belonged to lower socioeconomic status. While most of their parents were found to have limited education, they were considerate of their kid”s health. None of the children enrolled was found malnourished.

With regards to mean±SD serum 25-hydroxycholecalciferol levels, these were found to be significantly lower in children with ASD (9.53±4.93 IU/ml) and ID (14.39±5.99 IU/ml) compared to healthy controls (16.37±10.68 IU/ml) [F-value=3.61; P=0.032]. However, when utilizing standard cutoffs for vitamin D deficiency or insufficiency, we found that a non-significantly higher number of cases of ID and ASD were either Vitamin D3 deficient or insufficient, compared to the control group (P=0.076) [**Table 2**]. On detailed history-taking, it was found that only five parents from the control group were providing their kids with vitamin D3 supplementation; which also was infrequent and/or inadequate. Comparing the odds of ASD versus controls in those with vitamin D deficiency and those without, the odds ratio was found to be 6.0 (95% CI 0.68 to 52.75). All the patients were provided with serological reports and vitamin D3 supplementation was advised. The majority of children with ASD or ID and healthy controls had normal TSH levels, with only 8% (N=6/75) having raised levels [**Table 2**].

**Table 2:**
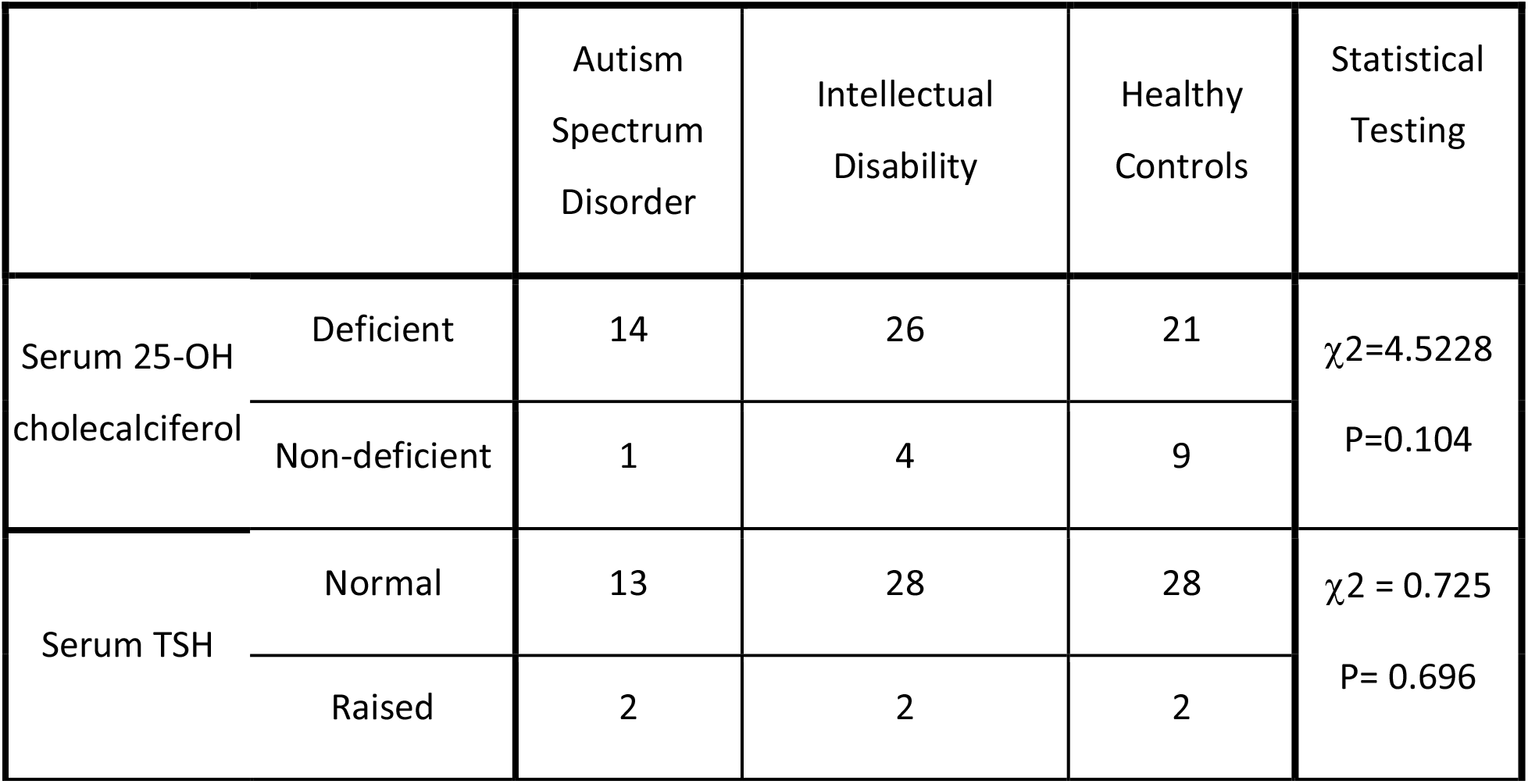
Comparison of Serum Vitamin deficiency levels. Applied χ2 test for significance.

Mean±SD levels of TSH were similar in ASD (2.65±1.96), ID (2.47±2.03) and controls (2.19±1.42), with no significant difference [F-value=0.37, P=0.69; **Table 3**].

**Table 3:** Comparison of Serum TSH and Vitamin D3 mean levels in study groups. Applied one way ANOVA for significance.

| Variable | Autism Spectrum Disorder |  | Intellectual Disability |  | Normal |  | F value (ANOVA) | P-value |
| --- | --- | --- | --- | --- | --- | --- | --- | --- |
|  | Mean | SD | Mean | SD | Mean | SD |  |  |
| Vitamin D level | 9.53 | 4.93 | 14.39 | 5.99 | 16.37 | 10.68 | 3.61 | **0.032** |
| TSH level | 2.65 | 1.96 | 2.47 | 2.03 | 2.19 | 1.42 | 0.37 | 0.691 |

## 4. DISCUSSION

### 4.1 Principal Findings

Literature regarding the role of endocrinology in ASD and ID, particularly from LMICs remains inadequate. Little data exists from LMIC settings investigating the aberrations in vitamin D and TSH levels in ASD and ID, like the present work. This study found a significantly lower serum 25-hydroxycholecalciferol in children with autism spectrum disorder (ASD) and intellectual disability (ID), compared to age-matched healthy controls, in a small sample from a resource-limited setting from an LMIC. These findings are consistent with prior literature, as discussed below.

The prevalence of overt and subclinical hypothyroidism has been reported to be 0.4% and 6.1% respectively by Marwaha, et al. amongst children in India (Marwaha et al., 2012). Our data indicate a hypothyroidism rate of 8% (N=6/75) in the total study population, being 13.3% in ASD, 6.7% in ID, and 6.7% in healthy controls. The abnormalities seen to be associated with hypothyroidism in the pediatric population are weight gain, increased cholesterol levels, impaired growth velocity, anemia, sleepiness, weakness, and impaired psychomotor and cognitive development (Aijaz et al., 2006). Given we did not have overt symptoms in any of the included cases, all individuals in our study with raised TSH were of subclinical hypothyroidism.

### 4.2 Comparison with Prior Literature

Wang et al. recently reported their findings from a large systematic review of the association between vitamin D and ASD. Pooling data from 20,580 participants, they found, similar to ours, lower levels of vitamin D concentration in cases of ASD than in controls (mean difference 7.46 ng/mL) (Wang et al, 2012). Low vitamin D levels were associated with a higher risk of ASD with an OR of 5.23 (95%CI 3.13 to 8.73), similar to our estimated OR value of 6.0. They also found that children with reduced maternal or neonatal vitamin D levels had an OR of 1.54 (95%CI 1.12 to 2.10) for developing ASD. Another recent meta-analysis pooled data from 5 RCTs of vitamin D supplementation for children with ASD (Li et al, 2022)

Mostafa and Al-Ayadhi studied vitamin D3 levels in autistic children aged 5-12 years and compared the results with age-matched controls. Similar to our observation they also observed significantly low levels of Vitamin D3 among autistic children (P < 0.001) with nearly 40% being vitamin D deficient (Mostafa and Al-Ayadhi, 2012). Similar observations were reported by Feng et al in 2016 from China, along with Meguid et al. in 2010 and Saad et al. in 2015, both from Egypt. The latter also conducted an open-label trial of vitamin D3 supplementation with 83 ASD children completing three months of 300 IU/kg/day of oral vitamin D3 and found over 80% of subjects had improved outcomes.

Mostafa and Al-Ayadhi also found that nearly 70% of their autistic patients had Increased levels of serum auto-antibodies of anti-myelin-associated glycoprotein (anti-MAG) type, whose levels were reported to negatively correlate with serum 25-hydroxy vitamin D levels. Frighi et al found that vitamin D deficiency rates were 77.0% in 155 ID patients and 39.6% in 192 controls (P<0.0001) from a cohort in the UK (Frigi et al., 2016). They also reported that winter season, darker pigmentation, reduced mobility, and obesity are associated with lower vitamin D levels; and recommended focused screening and specific therapeutic strategies for fracture risk reduction. Taken together, the literature supports the role of vitamin D deficiency in ASD and its potential as a therapeutic target.

The few studies that do not agree with the findings in the present work carry significant limitations. Hashemzadeh et al. (2015) did not find an association between vitamin D and ASD in children aged 3–12 years. However, they had a very low sample size [ASD (n = 13) and control (n = 14)] (Hashemzadeh et al., 2015). Another work, a conference paper by Basheer et al. which is yet unpublished, did not show an association. The authors themselves attributed this to the high prevalence of vitamin D deficiency in their population (Basheer et al., 2016).

Our work could not detect any specific association between serum TSH levels and ASD or ID. In the case of thyroid hormones, the literature is less clear compared to the consensus seen in the literature pertaining to vitamin D, especially from LMICs. To the best of our knowledge, there currently do not exist any studies from India examining the association between thyroid hormone aberration and ASD or ID. Desoky et al., through a case-control study, reported that where serum free T3 and free T4, were similar between 60 autistic and 40 control children, however serum TSH levels were higher in ASD, indicating the presence of subclinical hypothyroidism in children with autism (Desoky et al., 2017).

Hoshiko et al. reported an association between low thyroid hormones at birth and the occurrence of subsequently diagnosed ASD. They stated that hormone levels may return to normal or near-normal levels later but persistent neurodevelopmental impairment may exist (Hoshiko et al., 2011). This may explain why an association between hypothyroidism and ASD or ID was not detected in our work. A recent meta-analysis by Thompson et al. demonstrated that maternal subclinical hypothyroidism and hypothyroxinemia are associated with indicators of intellectual disability in the offspring but their effect on the risk of ASD in offspring was unclear (Thompson et al., 2018).

Findings from this work have further research and clinical Implications, particularly in the domain of Vitamin D deficiency and its correction thereof, in children with autism and ID. Targeted screening and risk factor mitigation solutions should be offered to these populations, both in LMICs and HICs. In addition, further high-quality randomized interventional studies are needed to assess whether vitamin D supplementation in early childhood of individuals with ASD may help improve outcomes later in life.

### 4.3 Limitations

Several limitations restrict the conclusions from the present. First, the sample size was small, arbitrary, and underpowered to detect subtle differences. Second, we did not capture data on several confounders. For instance, sunlight exposure, a known confounder, was not explored since it was felt that parent-reported estimates were unreliable in our set-up. Third, we did not utilize the severity of ASD and ID in our work which prevented us from exploring dose-response relationships. Fourth, the case-control nature of our work prevents any direct conclusions regarding causality. However, the resource-limited setting hindered the conduction of a prospective cohort study.

### 4.3 Conclusions

This study found significantly lower 25-hydroxycholecalciferol levels in children and early adolescents with autism spectrum disorder (ASD) and intellectual disability (ID) in Northern India. Given that deficiency of vitamin D has been implicated in their respective etiologies, high-quality randomized controlled trials are warranted to explore the therapeutic impact of early-life vitamin D supplementation in these indications. The utility of targeted screening and risk factor mitigation solutions may be explored in ameliorating these risk factors in high-risk populations across LMICs and HICs.

## Data Availability

All data produced in the present study are available upon reasonable request to the authors.

## Acknowledgments

We wish to acknowledge the contributions of the staff of the Chemical Pathology laboratory, the Department of Psychiatry, and the Department of Pediatrics.

## Prior publication or presentation

None

